# Compensatory mechanisms enables intelligibility in prodromal Parkinson’s disease

**DOI:** 10.1101/2022.12.14.22283451

**Authors:** Tabea Thies, Doris Mücke, Nuria Geerts, Aline Seger, Gereon R. Fink, Michael T. Barbe, Michael Sommerauer

**Affiliations:** University of Cologne, Faculty of Medicine and University Hospital Cologne, Department of Neurology, University of Cologne, Faculty of Arts and Humanities, IfL Phonetics; University of Cologne, Faculty of Arts and Humanities, IfL Phonetics; University of Cologne, Faculty of Medicine and University Hospital Cologne, Department of Neurology; University of Cologne, Faculty of Medicine and University Hospital Cologne, Department of Neurology, Institute of Neuroscience and Medicine (INM-3), Forschungszentrum Juelich

## Abstract

**Background:** Speech impairment is already present on the acoustic level in speakers with isolated rapid eye movement sleep behavior disorder (iRBD). The aim of this study was to determine whether speech changes are already present on the articulatory level and if how these differ from healthy control speakers and speakers with Parkinson’s disease (PD).

**Methods:** Kinematic data were collected from 68 age and sex-matched subjects: healthy control speakers (n=23), patients with iRBD (n=22), and patients with PD (n=23). All participants were recorded with electromagnetic articulography (AG 501) to capture articulatory movements of the lower lip, the tongue tip and the tongue body. Movement amplitudes, durations and average speeds were calculated per articulator. In addition, naïve listeners rated the intelligibility of the speech sampled produced by the participants.

**Results:** The results of the production experiment indicate changes between the control and the iRBD group as well as between the iRBD and the PD group. Movement durations increase from the control group to the iRBD group and further to the PD group. In contrary, movement amplitudes increase in prodromal PD and decreased from PD onwards. This relationship was reflected by preserved articulatory speed in iRBD patients, but articulatory slowdown in PD. Intelligibility is lower in speakers with PD, but does not differ between the control and the iRBD group.

**Conclusion:** Speakers with iRBD adjust underlying articulatory movement patterns to maintain their intelligibility level.

## Background

Patients with isolated rapid eye movement sleep behavior disorder (iRBD) are at high risk of developing Parkinson’s disease (PD). Furthermore, iRBD constitutes the prodromal marker with highest likelihood to develop PD^1,2^. Therefore, deviations in the speech motor system might evolve in patients with iRBD. Speech changes should develop in the direction of hypokinetic dysarthria which is characteristic in PD^3^.

Previous studies investigated the speech production in iRBD patients on the acoustic level. A reduced ability to modulate pitch was observed in speakers with iRBD^4–6^: Besides monopitch, articulatory deficits were reported most dominant ^7^ leading to a reduction of the acoustic vowel space was determined ^8^ as well as a trend towards lower articulation rates^5^. While acoustic speech parameters deviated between speakers with iRBD and a control group, no perceptual differences were identified by listeners^5^.

The aim of the present study was to directly track speech movements with electromagnetic articulography of patients with iRBD to investigate early articulatory alterations in prodromal PD. This enables us to go beyond the acoustic surface by tracking the underlying movement patterns of the articulators such as lips and the tongue system. As the acoustic vowel space and articulation rate were previously shown to be reduced in iRBD, smaller movement amplitudes (reduction in range) and longer movement durations (more time needed to achieve articulatory target) were expected. Data of speakers with iRBD were compared to speech patterns of age-matched healthy control speakers and speakers with PD.

## Methods

The study consisted of a production and a perception experiment. For the speech production part, 68 speakers were included in the study: 22 speakers with iRBD, 23 speakers with PD, and 23 healthy control speakers (Table 1). None of the participants showed signs of dementia or depression according to screening tests (Parkinson Neuropsychometric Dementia Assessment ^9^, cut-off < 15, and Beck’s Depression Inventory-II^10^, cut-off < 20). All participants gave consent prior to study inclusion. The study was approved by the ethics committee of the University of Cologne (18-425).

**Table 1:**
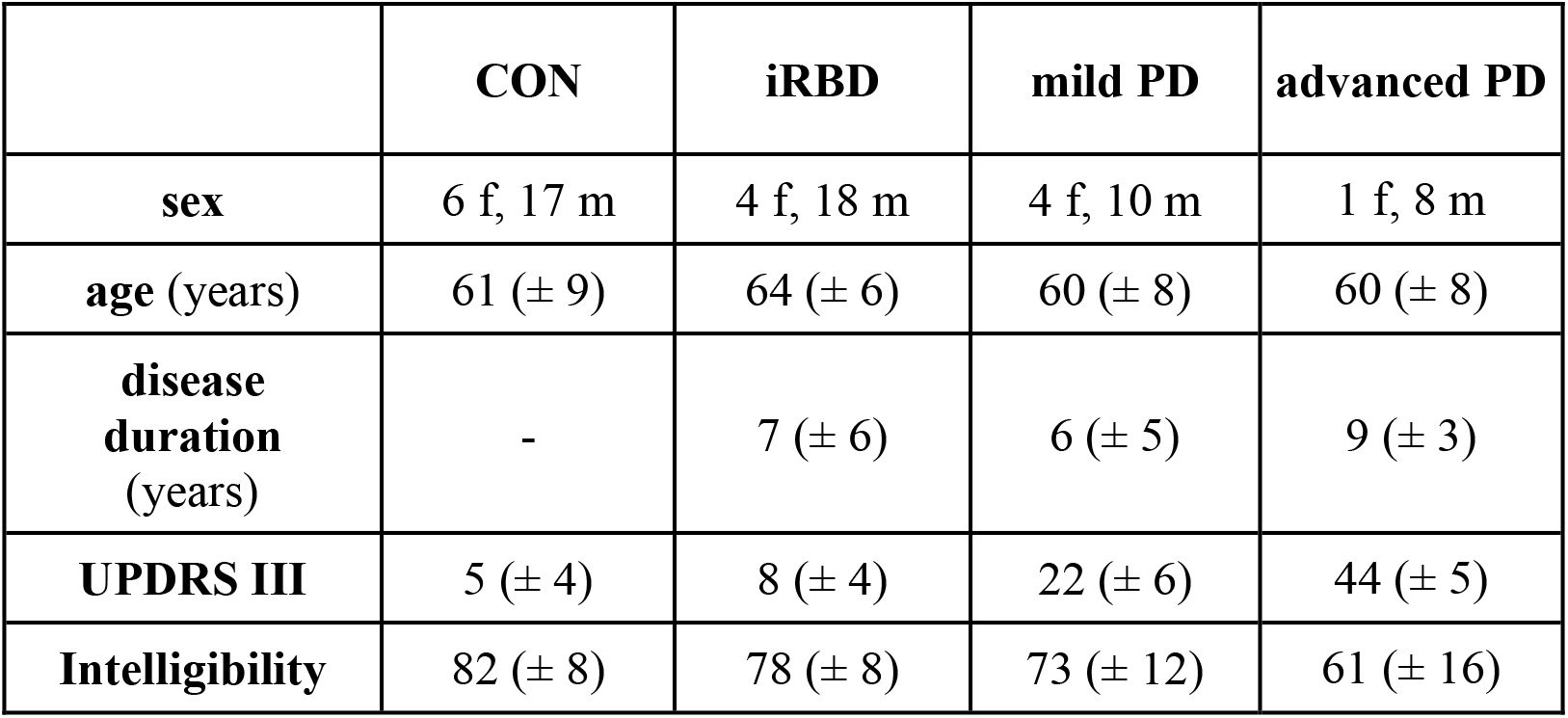
Demographics, gross motor scores and intelligibility ratings per group.

|  | **CON** | **iRBD** | **mild PD** | **advanced PD** |
| --- | --- | --- | --- | --- |
| **sex** | 6 f, 17 m | 4 f, 18 m | 4 f, 10 m | 1 f, 8 m |
| **age (years)** | 61 (± 9) | 64 (± 6) | 60 (± 8) | 60 (± 8) |
| **disease duration (years)** | - | 7 (± 6) | 6 (± 5) | 9 (± 3) |
| **UPDRS III** | 5 (± 4) | 8 (± 4) | 22 (± 6) | 44 (± 5) |
| **Intelligibility** | 82 (± 8) | 78 (± 8) | 73 (± 12) | 61 (± 16) |

Motor functions of all participants were assessed by using part III of the Unified Parkinson’s Disease Rating Scale (UPDRS)^11^. Gross motor and speech motor performance of patients with PD were assessed in a practical defined medication-OFF condition, after withdrawing PD medication for at least 12 hours, to capture the disease status reducing influence of PD drugs. The PD group was divided into two groups: one with mild motor symptoms (UPDRS III ≤ 32) and the other with more severe motor symptoms (UPDRS III ≥ 33)^12^.

Speech production data were elicited by using electromagnetic articulography (AG 501, Carstens). Small sensors were attached to the lower lip, the tongue tip and the tongue body to capture the spatio-temporal characteristics of articulatory movements during speech (Figure S1). The experimental set-up, the speech material and the data processing were the same as already described^13,14^. Speakers produced sentences, such as “Die Oma hat der Mila gewunken” (The grandma waved at Mila.). The focused element of the analysis was the first syllable from each of the ten girl names (Table S1), which were produced in three different focus conditions that reflect different degrees of prominence, ranging from low to high prominence on the name^15,16^. Tongue body movements refer to the production of the vowels (i, e, a, o, u), tongue tip movements to the production of the alveolar consonant /l/ and lower lip movements to the production of the labial consonant /m/. The articulatory movements producing these sounds were the unit of interest. For each articulatory movement, the amplitude (in mm) and the duration (in ms) were calculated (Figure S2). While the duration indicates how much time a speech movement takes, the amplitude refers to the spatial distance the articulator travels during this time interval. Furthermore, the average movement speed was calculated as a ratio of amplitude over duration.

In a speech perception experiment, 280 naïve listeners independently rated the intelligibility of a subset of sentences collected in the production experiment. The sentences that were included were produced in the most natural prominence condition (broad focus) and contained only girl names that have the vowels /i, e, u/ in their first syllable to reduce the time of the experiment and to counteract fatigue effects. Each sentence was at least rated by 40 raters. Ratings were elicited on a visual analogue scale, ranging from 1 to 101. Higher values indicate more intelligible speech.

Statistical analyses were performed with the software R^17^. Linear mixed models were applied using the ‘lme4’ package^18^ to test group effects on articulatory parameters. While group and prominence condition were predictor variables, random intercepts were included for speaker and vowel type as well as varying slopes for speaker per linguistic prominence condition. A random factor for consonant was added when investigating tongue body movement patterns only. Post-hoc analyses for pairwise group comparisons were conducted by using the ‘emmeans’ package^19^. Reported p-values are based on these analyses. Continuous ordinal regression models were applied to test differences in intelligibility ratings across the groups^20^. Group, vowel and consonant were predictor variables. Random intercepts were included for speaker and rater. Significance was accepted at p < 0.05. The data that support the findings of this study are available from the corresponding author upon reasonable request.

## Results

Groups did not differ significantly regarding age, and sex. MDS-UPDRS III scores increased from healthy controls to patients with iRBD to mild PD and further to more advanced PD (Table 1). Intelligibility ratings decreased from healthy controls to mild PD (β = 0.87, p = .008) and further to advanced PD (β = 1.85, p = .001).

The results of the production experiment revealed that speakers with iRBD produced longer tongue tip movements (t(70.0) = -3.214, p = .01), larger tongue body (t(72.2) = -3.509, p = .004) and larger tongue tip movements (t(75.9) = 2.457, p = .02) compared to control speakers (Figure 1). Movement durations increased further from the iRBD group to the PD group, while movements amplitudes decreased. Changes in movement duration were seen for tongue tip (CON vs. mild PD: t(70.0) = -4.456, p < .001 | CON vs. advanced PD: t(70.8) = -4.030, p < .001) and lower lip movements (CON vs. mild PD: t (71.6) = -3.673, p = .003). Amplitudes reduced from iRBD to advanced PD also for tongue tip (iRBD vs. mild PD: t (61.9) = 2.707, p = .043 | iRBD vs. advanced PD: t(62.6) = 3.366, p = .007) and lower lip movements (iRBD vs advanced PD: t(73.1) = 3.068, p = .016). Furthermore, the results of the average speed of vocalic tongue body movements revealed a steadily increase from the control group to advanced PD because durational properties remained the same, while the amplitudes enhanced (Figure 1). In contrast, the average speed of consonantal movements of the tongue tip and the lower lip steadily decreased as movement duration increase while amplitudes reduce.

**Figure 1:**
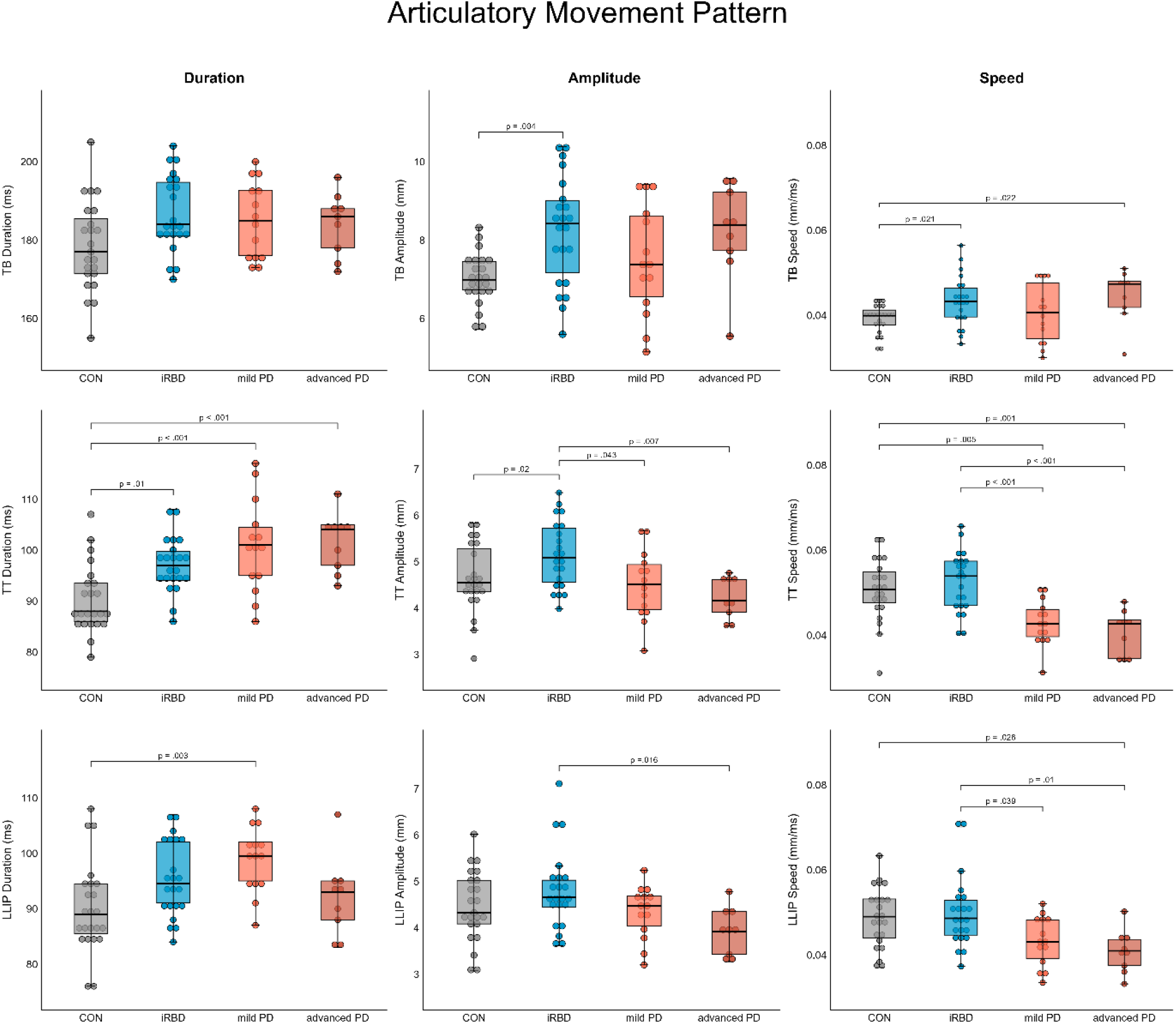
Articulatory results per group and parameter as averages across prominence conditions.

## Discussion

Although, patients with iRBD did not show gross motor symptoms, or a reduced intelligibility, compensatory changes on the underlying articulatory mechanisms were detected. The results of the production experiment showed changes between the control and iRBD group as well as between the iRBD and PD group (Figure 1). Movement durations became longer from the control group to the iRBD group and further prolongate in PD as soon as gross motor performance is more affected. In contrary, movement amplitudes were increased in prodromal PD and decreased from PD onward. This was reflected by preserved articulatory speed in iRBD patients, but articulatory slowdown in PD.

Articulatory movements of speakers with iRBD take more time to achieve the articulatory target, but contrary to the expectations of reduced movement ranges, larger tongue movement amplitudes were determined. Longer and larger movements are leading to a more distinct speech but they also require an increase in biomechanical effort of the articulators during speech (cf. hyper-articulation^24^). In general, speakers aim to produce speech by following the principle of physical economy, i.e., they balance the degree of speech intelligibility and the amount of articulatory costs.. However, it has been shown that speakers adapt their way to speak dependent on internal or external factors, such as physiological conditions. As it can be seen in this data set, the speech effort of tongue movements in particular increased in speakers with iRBD compared to control speakers.

The observed speech pattern in the iRBD group can be interpreted as a compensation mechanism to counteract effects of incipient motor detriment on speech to reach the articulatory goal. As usually smaller amplitudes lead to reduced intelligibility in dysarthric speech^25–27^, this compensatory mechanism allows to maintain the level of intelligibility as it does not differ from healthy control speakers. Furthermore, larger articulatory movements were investigated in mild-dysarthric speakers with PD before^28^. Thus, the hyper-articulation strategy may indicate that speakers with iRBD are developing a dysarthria which cannot yet be perceived auditorily, which is in line with results of a previous study. Moreover, subtle speech changes in the iRBD group are related to lingual involvement only.

Speakers with PD are less intelligible and show a slowdown of the tongue tip and the lower lip indicating that dysarthria has already further developed from lingual to labial involvement^29^. While amplitudes of vocalic movements further increase from mild PD to advanced PD, they decrease for consonantal movements. The reduction in amplitude is accompanied by a prolongation in the temporal domain and a decrease of the average speed. Knowing now that there may have been a phase of compensation (in terms of spatial enhancement) prior to the spatial reduction, speakers with PD do no longer compensate for disease effects on their speech by means of spatial adjustments. The compensation mechanism may no longer be used as soon as motor symptoms are pronounced. Thus, reduction in the movement amplitude of the tongue tip and lower lip might be a relevant indicator of a developing PD in speakers with iRBD.

## Data Availability

All data produced in the present study are available upon reasonable request to the authors.

