## Supplements for "Compensatory mechanisms enables intelligibility in prodromal Parkinson’s disease"

**Supplement:**

**Table S1:** Ten target words with varying consonant vowel sequence.

| vowel | labial C1 /m/ | alveolar C1 /l/ |
|---|---|---|
| /a/ | Mali | Lani |
| /e/ | Mela | Lena |
| /i/ | Mila | Lina |
| /o/ | Moli | Loni |
| /u/ | Mula | Luna |

**Figure S1:** Sensor positions in the mid-sagittal plane on the lips, tongue tip and tongue body.

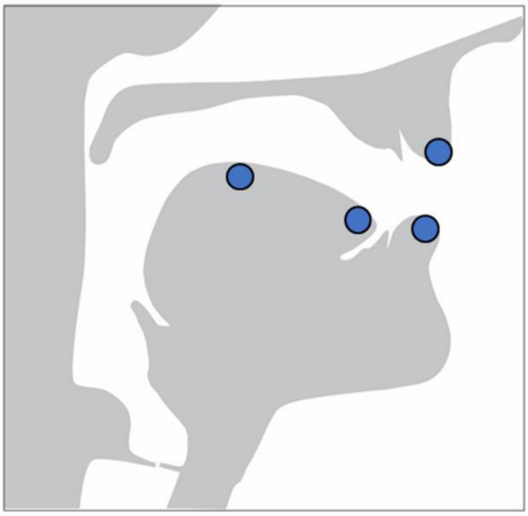

**Figure S2:** Schematized articulatory movement and related measures. Duration measured in milliseconds and amplitude in millimeters.

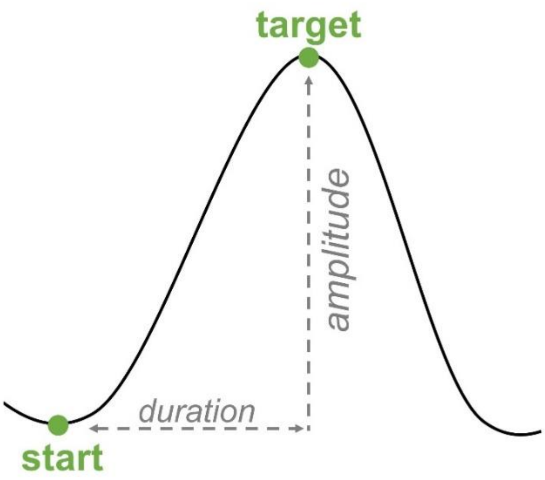
